# Use of Convalescent Plasma Therapy among Hospitalized Coronavirus Disease 2019 (COVID-19) Patients: A Single-Center Experience

**DOI:** 10.1101/2021.02.16.21251824

**Authors:** Flordeluna Z. Mesina, Claudette G. Mangahas, Ellen M. Gatchalian, Mary Sheila Ariola-Ramos, Rosalio P. Torres

**Affiliations:** Section of Hematology, Department of Medicine, Cardinal Santos Medical Center, San Juan, Metro Manila, Philippines; Section of Pulmonary Medicine, Department of Medicine, Cardinal Santos Medical Center, San Juan, Metro Manila, Philippines; Section of Infectious Disease, Department of Medicine, Cardinal Santos Medical Center, San Juan, Metro Manila, Philippines

**Keywords:** Convalescent plasma, COVID-19

## Abstract

COVID-19 Disease has strained our healthcare system. Convalescent plasma has been used to treat emerging infectious diseases -Influenza A/B, SARS-CoV, Ebola virus and now SARS-CoV 2.

**OBJECTIVE:** This study aims to determine the outcome and clinical course of COVID-19 patients who received convalescent plasma transfusion at Cardinal Santos Medical Center.

**METHODS:** This is a retrospective cohort analytical study of 75 patients who received convalescent plasma.

**RESULTS:** Median time from admission to CP transfusion was 3 days. Majority of patients received additional therapies including dexamethasone (100%); Remdesivir (95%); antibiotics (100%), tocilizumab (65%); hemoperfusion (88%) or combination of these. Among the survivors, the median LOS was 15 days while non-survivors have a median LOS of 6 days. One patient (1.33%) had mild transfusion reaction. Four patients (5.33%) developed DVT despite anti-coagulation. There was improvement in the inflammatory markers (LDH pvalue 0.04, CRP pvalue 0.00, Ferritin pvalue 0.0001). There was improvement in the pulmonary parameters -increase in mean PaO2, mean SaO2, and mean PFR; and decrease in mean FiO2 and mean RR post-treatment. Median LOS is 14 days for the CP group vs 11 days for the non-CP group. Mortality rate among the CP group is 25.33% while the non-CP group was 26.67%. LOS and mortality rate did not reach statistical significance.

**CONCLUSIONS:** There was no significant difference in mortality and length of hospital stay in patients given CP vs controls. CP might have a role in the improvement of inflammatory markers and pulmonary status.

## BACKGROUND

The Novel Coronavirus Disease 2019 (COVID-19) has gripped our country with overwhelming strain in our health system, while there is no single medication which offers cure, we would like to look into the possible benefits of convalescent plasma (CP) in limiting complications and in treating COVID-19. Plasma is the liquid portion of blood containing proteins like albumin, coagulation cascade factors, complement, and a variety of antibodies or immunoglobulins and enzymes.^1^ The primary function of all immunoglobulins is the recognition and binding of specific antigenic determinants, whether soluble (including toxins), particulate, or cellular (such as pathogens). The consequences of immunoglobulin binding depend on the nature of the antigen, and at its simplest, may be the physical prevention of antigen penetration through the epithelium.^2^ This is the neutralizing effect of immunoglobulins. Secondary effects of immunoglobulin are complement activation, opsonization and antibody dependent cell mediated toxicity by different cells.

The premise behind use of convalescent plasma in COVID-19 patients is the passive transfer of antibodies from recovered individuals to patients with the virus manifesting with diseases. There is both historical precedence and biological plausibility for the use of CP as it has been utilized in previous pandemics. In the Spanish flu of 1918, there was a systematic review on the use of convalescent blood products which showed case fatality rate of 19% in patients given CP < 4days vs. 59% for those given > 4 days from onset of symptoms.^3^ In 1974, the Argentine hemorrhagic fever outbreak, there was a double blind placebo controlled study which showed 1.1% case fatality rate in those given immune plasma vs 16.5% in those given normal plasma. In 2011 during the Influenza A/B outbreak where patients were given CP + standard of care vs. standard of care alone, it showed a non-significant reduction in time to normalization of patients’ respiratory status; significant reduction in mortality was noted. It was also noted to be safe and well tolerated.^4^ The EBOLA outbreak in 2015 in west Africa, the study showed there was no difference in the risk of death at 31% in CP group vs 38% in the control group and that there were no adverse events noted.^5^ CP was also used in other coronavirus such as the SARS epidemic in 2003. A retrospective study in Hongkong showed good clinical outcome in 33 out of 80 patients and improved outcome was associated with early administration of CP.

The use of convalescent plasma in COVID-19 is still in the experimental stage and is not yet included as a standard of care for COVID-19. With the unequivocal efficacy data and good safety profile from published studies, we believe that more studies are needed. It has been utilized in our institution through the Hematology Section Convalescent Plasma Program as an adjunct treatment and it is our aim to collate, analyze and report data of patients who received CP.

### OBJECTIVES

#### Primary Objective

To determine the outcome and clinical course of COVID-19 patients who received convalescent plasma transfusion in our institution

#### Specific Objectives

1. To describe the clinical and laboratory profile of COVID-19 patients who received convalescent plasma transfusion
2. To determine effect of CP on pulmonary parameters and inflammatory markers pre and post CP transfusion
3. To determine the effect of convalescent plasma compared to historical controls (age, gender and severity-matched controls) with regards to:
  a. Length of hospital stay
  b. all-cause mortality
4. To determine incidence of adverse events of convalescent plasma transfusion among recipients.

## METHODOLOGY

### Research design

This is a retrospective cohort analytical study.

### Population and Sampling

#### Inclusion criteria

1. Patients must be 18 years of age or older
2. Hospitalized and diagnosed with COVID-19, confirmed by quantitative reverse-transcriptase polymerase chain reaction (QRT-PCR) for SARS COV-2 through nasopharyngeal/oropharyngeal swab (NPS/OPS)
3. Patients who are confirmed COVID-19 with moderate pneumonia, severe and critical were included in this study

### Controls/Non-CP Group

The patients who received CP were matched based on age, gender and severity of COVID-19 pneumonia to patients who were admitted from April 1, 2020 to October 31, 2020 who received other COVID-19 treatment except convalescent plasma. There were several reasons why these patients did not received convalescent plasma: 1) attending physicians discretion; 2) no available plasma with the same blood type; 3) they were admitted during the time where CP program was not yet available in our institution; OR 4) they were referred for CPT but did not give consent for plasma transfusion.

### Exclusion criteria

Patients with incomplete medical records/data were excluded.

### Sampling

The study used convenience sampling. All patients who received convalescent plasma from April 1, 2020 to October 31, 2020 were included.

### Data Collection

Investigators collected data using google forms and excel sheets. The following data were captured: demographic data (age, sex); medical data (diagnosis, date of admission, date of discharge, date of CP transfusion and day of illness, comorbidities, adverse events); treatment data (medications received; outcome). The sources of data were patient chart from the medical records section and/or census. Data custodian ensured completeness of data and consistency with source data.

### Data Analysis

Descriptive statistics using frequency, mean, median and mode were used. For the comparison of pulmonary and inflammatory parameters pre and post-CP transfusion, Wilcoxon signed rank test was used. For the comparison between CP and non-CP group with mortality and length of hospital stay, Mann Whitney U test was used.

### Ethical Considerations

The study proposal was reviewed and approved by the CSMC Research Ethics Review Committee.

## RESULTS

There were seventy five (75) COVID-19 patients diagnosed using QRT-PCR for SARS COV-2 on NPS/OPS given convalescent plasma from April 1, 2020 to Oct 31, 2020 who were included in this retrospective analysis. One patient was excluded because of incomplete data and death in <48 hours. Demographics and baseline data of the participants are shown in Table 1. The median age is 65 years old (range of 30 to 88 years old), majority 39/75 (52%) are 51 to 70 years old and 23/75 (30.67%) are 71 to 90 years old. That makes a combined proportion of 82.67% belonging to the >50 years old age group. Forty-five (60%) are males and 30 (40%) are females. With regards to COVID-19 pneumonia severity, most of the patients referred for convalescent plasma were severe 32 (42.67%) and critical 23 (30.67%). Majority of the patients (93.33%) required some form of oxygen support in the form of nasal cannula, face mask, high flow nasal cannula or mechanical ventilation.

**Table 1.** Characteristics of CP group.

| FEATURE | N=75 (%) |
| --- | --- |
| Age |  |
| 18-30 | 1 (1.33%) |
| 31-50 | 12 (16%) |
| 51-70 | 39 (52%) |
| 71-90 | 23 (30.67%) |
| >90 | 0 |
| All (median) | 65 years old (100%) |
| Sex |  |
| Female | 30 (45%) |
| Male | 45 (65%) |
| COVID pneumonia severity classification |  |
| Moderate | 20 (26.6%) |
| Severe | 32 (42.67%) |
| Critical | 23 (30.67%) |
| Type of Oxygen support |  |
| Room Air | 5 (6.67%) |
| Nasal Cannula/Face mask | 25 (33.33%) |
| HFNC | 25 (33.33%) |
| Intubated | 20 (26.67%) |
| Blood type |  |
| A+ | 11 (14.67%) |
| B+ | 24 (32%) |
| O+ | 33 (44%) |
| AB+ | 7 (9.33%) |
| Baseline laboratories |  |
| Hemoglobin |  |
| Hb < 12 g/dl | 20 (26.67%) |
| Hb 12-18.0 g/dl | 55 (73.33%) |
| Hb > 18.0 g/dl | 0 |
| WBC |  |
| WBC < 5.0 | 11 (1.33%) |
| Normal 5-10 | 37 (49.33%) |
| WBC > 10.0 | 27 (36%) |
| Neutrophils | 0.81 (0.53-0.96) |
| Lymphocytes | 0.13 (0.03-0.85) |
| NL Ratio |  |
| Normal (1-3) | 9 (12%) |
| Abnormal NLR (>3) | 66 (88%) |
| Platelet |  |
| < 150,000 | 14 ( 18.67%) |
| 150,000-450,000 | 61 (81.33%) |
| >450,000 | 0 |
| CRP | 206.02 (3.9-6144) SD 700.57 |
| LDH | 485.82 (138-1347) SD 247.31 |
| Ferritin | 3480.41 (444-61,256) SD 7347.79 |
| D-dimer (median) | 1380 (24->5000) |

Baseline laboratories showed that the majority of patients had a hemoglobin of 12-18 g/dL accounting for 55 (73.33%) while 20 (26.67%) had low hemoglobin. Leukopenia was seen in 11 (1.33%) of patients while leukocytosis in 27 (36%). The mean neutrophil count is 0.80 (range 0.53 to 0.96) and mean lymphocyte count is 0.13 (range 0.03 to 0.85). Neutrophil-lymphocyte ratio (NLR), a marker of inflammation, was computed and showed that majority had an abnormal NLR of above 3 accounting for 66 (88%). Majority of the patients, 61 (81.33%) had normal platelet count while only 14 (18.87%) had thrombocytopenia at baseline. All of the patients had elevated inflammatory markers. The mean lactate dehydrogenase (LDH) was 485.82 (138 to 1347); mean serum ferritin was 3480.41 (444 to 61,256); mean C-reactive protein (CRP) was 206.02 (3.9 to 6144) and median D-dimer was 1380 (24 to >5000).

The median time from hospitalization/admission to CP transfusion was early at 3 days, most of them given within 2–5 day from admission (interquartile range [IQR], 2-5 days). The majority of patients received additional therapies including dexamethasone (100%); Remdesivir (95%); all were given antibiotics either piperacillin-tazobactam or meropenem or ceftriaxone and/or azithromycin, tocilizumab (65%); hemoperfusion (88%) or multiple combinations of these. CP recipients had primarily O+ blood type (44%) followed by B+ at 32%.

The outcomes of patients are summarized in Table 2. The median length of stay (LOS) of all patients was 14 days [IQR 9-20 days]. Among the survivors, the median LOS was 15 days [IQR 10-20 days] while it is shorter in non-survivors with a median LOS of 6 days [IQR 6-15.5 days]. One patient (1.33%) had mild transfusion reaction which was managed with intravenous antihistamine. Four patients (5.33%) developed DVT during the course of hospitalization despite anti-coagulation. There were 19/75 patients (25.33%) who died from the CP recipients. The most common cause of death is septic shock secondary to pneumonia.

**Table 2:** Course and Outcome of Patients given CP.

| Parameters | Results |
| --- | --- |
| Timing of CP transfusion from admission, median [IQR] | 3 days (2-5 days) |
| <u>Length of Hospital Stay (LOS)</u><br>LOS of survivors, median [IQR]<br>LOS of non-survivors, median [IQR]<br>Overall LOS, median [IQR] | 15 days (10-20 days)<br>6 days(6-15.5 days)<br>14 days (9-20 days) |
| <u>Adverse Events</u><br>Transfusion reaction<br>Thrombosis(acute DVT or stroke)<br>Total | 1/75 (1.33%)<br>4/75 (5.33%)<br>5/75 (6.67%) |
| <u>Outcome</u><br>Dead<br>Alive | 19 (25.33%)<br>56 (74.66%) |

### PRE and POST TREATMENT DATA

Laboratory parameters were collected pre-treatment which was either laboratory results during admission or the nearest prior to CP transfusion and post-treatment which was within 7 days after CP transfusion shown in Table 3. There was a decrease in the mean hemoglobin at 13.24 g/dL pre-treatment to 12.90 g/dL post-treatment. There was an increase in mean WBC from 10.00 to 11.60 pre– and post–treatment. These were statistically significant. There was no significant change in the mean platelet level from 235.60 to 251.20, very few had mild thrombocytopenia.

**Table 3:** Laboratory and Pulmonary Parameters Pre- and Post-treatment.

| Parameters | Pre treatment | Post Treatment | p-value |
| --- | --- | --- | --- |
| Hemoglobin (g/dl) mean | 13.24 (8.1-18) | 12.90 (8.20-20) | 0.005 |
| WBC (X 10<sup>9</sup>/L) | 10.00 (1.75-31.13) | 11.60 (2.22-31.68) | 0.003 |
| Neutrophils (%) | 80.85 % (53%-96%) | 80.95% (7%-97%) | 0.45 |
| Lymphocytes (%) | 13.05 % (3-85%) | 11.24 (1-31%) | 0.27 |
| NL Ratio | 10.30 (1.04-31.67) | 12.31 (1-97) | 0.41 |
| Platelet (X 10<sup>9</sup>/L) | 235.60 (53-417) | 251.20 (28-664) | 0.12 |
| LDH (U/L) | 485.82 (138-1347) | 453.24 (48-1395) | 0.04 |
| CRP (U/L) | 206.02 (3.9-6144) | 53.98 (2.7-330) | 0.0000 |
| Ferritin (mg/dl) | 3480.41 (444-61,256) | 2713.54 (125-33,781) | 0.0001 |
| PaO<sub>2</sub> | 90.6 (40.9-355) | 91.73 (42-272) | 0.58 |
| SaO<sub>2</sub> | 94.20 (84.1-100) | 95.35 (81-100) | 0.014 |
| FiO<sub>2</sub> | 54.21 (21-100) | 49.25 (21-100) | 0.0084 |
| PFR | 193.29 (48-461) | 231.72 (47-461) | 0.0007 |
| RR | 24.28 (18-40) | 22.15 (16-38) | 0.0000 |
| ROX index* | 9.04 (3.62-18) | 12.78 (2.84-22.6) | 0.0000 |
| **Chest Imaging (X-ray/CT):** |  |  |  |
| Improved |  | 45/75 (60%) |  |
| Stable/No change |  | 8/75 (10.67%) |  |
| Progression |  | 19/75 (25.3%) |  |
| No repeat Chest Radiograph |  | 3/75 (4%) |  |

*\*computed only for selected patients*

There was a noted trend towards improvement in the inflammatory markers, which were statistically significant. The mean LDH from a baseline was 485.82, which decreased to 453.24 ng/mL, mean pre-treatment CRP was 206.02 which decreased to mean post-treatment of 53.98 and mean serum ferritin also decreased from 3480.41 to 2713.54. These were all statistically significant.

Pulmonary parameters which denoted oxygenation status also trend towards improvement as there was an increase in mean PaO2, increase in mean SaO2, decrease mean FiO2, increase in mean PFR and decrease mean RR post-treatment. ROX index was computed for selected patients on high flow nasal cannula which showed a significant improvement in the mean ROX index pre and post treatment. Chest imaging improvement pre-treatment and post-treatment were noted in 45/75 (60%) of patients; 19/75 (25.3%) progression of chest findings; 8/75 (10.67%) no change and 3/75 (4%) had no repeat imaging.

### Comparison of CP group vs matched-controls

Patients given CP were matched by age, gender and severity of illness with control patients who were given other forms of treatment except convalescent plasma. Table 4 summarizes the comparison between the two groups with regards to the length of hospital stay and mortality. Among the survivors from the CP group, the median LOS is 15 days with most of them staying in the range of 10 to 20 days while the median LOS among the non-CP group is 11.5 days with an IQR of 9-17 days. Among the non-survivors from the CP group, the median LOS is 6 days with an IQR of 6 to 15.5 days, while the median LOS among the Non-CP group non-survivors is 11 days with an IQR of 6.5 to 26 days. The median LOS of all patients is 14 days with an IQR of 9 to 20 days for the CP group vs 11 days for the non-CP group with an IQR of 8 to 17 days. All values when compared did not reach statistical significance.

**Table 4:** Comparison of Length of hospital stay and mortality CP vs non-CP historical controls.

| Parameter | CP | Non-CP | p-value |
| --- | --- | --- | --- |
| <u>Length of Hospital Stay (LOS)</u> |  |  |  |
| LOS of survivors, median [IQR] | 15 days (10-20 days) | 11.5 days (9-17 days) | 0.25 |
| LOS of non-survivors, median [IQR] | 6 days(6-15.5 days) | 11 days (6.5-26 days) | 0.71 |
| Overall LOS, median [IQR] | 14 days (9-20 days) | 11 days (8-17 days) | 0.19 |
| All-cause mortality (n;%) | 19/75 (25.33%) | 20/75 (26.67%) | 0.85 |

The mortality rate among the CP group is 25.33% (19/75) while the non-CP group was 26.67% (20/75) and was not statistically different.

## DISCUSSION

This study reports our experience on the use of convalescent plasma in COVID-19 not just in severe and critical patients but also COVID-19 with moderate pneumonia. Our patient profile consisted of the majority elderly wherein 82.67% belonged to the >50 years old age group and male gender making up 60% of the subjects. This is consistent with the epidemiologic study of Haw et al. that COVID-19 in the Philippines is more common in males and older age groups.^6^ Genetics, behavior/lifestyle and immune system were factors cited for this biological differences.^7^ Hematologic profile showed anemia which may be due to inflammation and/or bleeding during the course of hospitalization. Leukopenia common in viral infection, was less frequently seen in our study. Normal to mild thrombocytopenia was frequent. The NLR which is a marker of inflammation was high in 88% of the subjects. There were many studies showing a vast range of hematological abnormalities in COVID-19 patients, the most common findings include lymphocytopenia, neutrophilia, eosinopenia, and mild thrombocytopenia.^8-11^ According to a meta-analysis, increased white blood cell count, and decreased lymphocyte and decrease platelet counts are associated with severe disease and higher mortality.^12^ A study by Ellinghaus et al., showed that “O” blood type was associated with lower risk of infection compared to non “O” blood type, while “A” blood type had greater risk than non A persons.^13^ In our study, though we did not look into the association of blood groups to severity of disease, it was noted that most of our COVID patients are O+ at 44% followed by B+ at 32%. This may be due to the fact that most Filipinos have O+ blood type at 44 to 46%.^14^

We looked into mortality, length of hospital stay, laboratory and clinical parameters pre and post treatment as measures of effectiveness of CP treatment. The results showed all-cause mortality and length of hospital stay were not statistically significant between CP and controls, at 25.33% vs 26.67% (p value 0.85) and 14 days vs 11 days (p value 0.19), respectively.

There were several non-randomized studies and observational studies and RCTs with conflicting data regarding the efficacy of CP. The following related studies showed results favoring CP. Shen et. al was a case series of 5 critically ill COVID-19 patients given CP which showed improvement in fever, SOFA score, P/F ratio, and decrease viral load. Four patients recovered from acute respiratory disease syndrome (ARDS), and three were weaned from mechanical ventilation within 2□weeks of treatment.^15^ Duan et al. reported 10 critically ill COVID patients who were given CP and compared them with control group. This study showed clinical, laboratory and radiologic improvement in those given CP, mortality rate was 0 in CP vs 3/10 in the historical controls.^16^ An RCT from Spain involving 81 patients showed improved survival in the convalescent plasma group (mortality at 15 and 29 days, 0% with convalescent plasma and 9% [4 of 43] in controls. The likelihood of progression was also lower with convalescent plasma (0% with convalescent plasma versus 14% of controls).^17^ Li and Zhang et al study was an RCT from china involving 103 patients who received CP vs standard of care, mortality rate was 16% vs 24%; time to improvement was 2 days vs 5.3 days.^18^ Rasheed et al. a multicenter study in Iraq with a population of 49 comparing those who were given CP (21) vs best available treatment (BAT) (28). The results showed a mortality of 1/21 (5%) vs 8/28 (29%) and a shorter length of hospital stay. ^19^ The biggest study so far was from the Mayo clinic collaboration in USA with a total of 35,322 subjects which showed transfusions of CP within 3 days of disease yielded greater reduction in mortality that is 8.7% vs 11.9% in late CP; a higher antibody titer was also associated with lower mortality.^20^ There were also negative studies, Zeng et al. reported 6 critically ill COVID-19 patients which showed no significant difference in mortality in those who received CP (5/6 patients) vs control group (14/16 patients).^21^ Salesi et al. is a multicenter study from Iran with a recruited population of 189. It compared hospitalized COVID-19 patients who were given CP (115) vs BAT (74). Results showed a non-significant reduction in mortality that is 14.8% vs 24.3% (p=0.09) and shorter length of hospital stay 9 days vs 13 days (p=0.002).^22^ The PLACID trial is the first RCT with a large population involving 464 patients from India which showed CP was not associated with a reduction in the progression from moderate to severe covid-19 and no reduction in all-cause mortality.^23^ And finally, a recent living systematic review by Piechotta et al. showed uncertain effect on mortality with a risk ratio of 0.89 (CI 0.61 to 1.31).^24^

With regard to safety of CP, published data are consistent with the low incidence of adverse events. Our study showed a mild infusion reaction of 1/75 (1.33%) and acute DVT of 4/75 (5.33%). Joyner et al. in a multicenter collaborative study of 20,000 hospitalized patients who received CP reported <1% transfusion reaction, <1% thromboembolic events and 3% cardiac events.^25^ Most of the studies mentioned above also noted none to low incidence (<1%) of adverse events after CP transfusion.

Although it seems that overall outcome in terms of mortality and length of hospital stay did not significantly impact those who received CP compared to controls, our study showed that there was a trend in the improvement of inflammatory markers, pulmonary parameters and oxygenation status pre and post treatment in the majority of patients and these were statistically significant. These findings were also reported in many case series and observational studies. An early study in china reported 4 patients recovered from ARDS and 3 weaned from mechanical ventilator within 2 weeks of CP administration.^15^ Duan’s pilot study showed laboratory, clinical and radiologic improvement in 10 critically ill covid-19 patients.^16^ A case series from South Korea, reported improvement in 3 COVID-19 patients with ARDS on mechanical ventilator.^26^ Eight out of 10 critically ill COVID-19 patients from Mexico and 18 out of 20 patients from a US study reported improvement in inflammatory markers, clinical and radiologic improvement.^27-28^ Whether it is due to convalescent plasma and/or other therapies given to patient, our study is not designed to determine this.

One limitation of this study, is that antibody titer in the donors’ plasma was not quantitatively determined because of the unavailability of the test. It was seen in a large multi-center study by Joyner et al. that there’s a significant survival advantage in COVID-19 patients who were given high antibody titer plasma (>18.45 S/co) compared to low titer (<4.62 s/co).^20^ It might have contributed to the non-significant difference in the mortality rate and hospital stay seen in our study.

## CONCLUSION

In conclusion, although there was no difference in mortality and length of hospital stay in patients given CP vs controls. CP might have a role in the improvement of inflammatory markers and pulmonary status of COVID-19 patients. And finally, CP transfusion is relatively safe with low incidence of adverse events.

## Data Availability

Data referred in the manuscript are available.

## ACKNOWLEDGEMENTS

We are sincerely thankful to our colleagues who helped us in completing this endeavor: Dr. Arvin Jay Bautista, Dr. Erika Chua, Dr. Matt Fontarum, Dr. Elijah Adamos, Dr. Marge Mesina and Dr. Sharlene Dizon.

